# Assessing the Impact of Imputation on the Interpretations of Prediction Models: A Case Study on Mortality Prediction for Patients with Acute Myocardial Infarction

**DOI:** 10.1101/2020.06.06.20124347

**Authors:** Seyedeh Neelufar Payrovnaziri, Aiwen Xing, Shaeke Salman, Xiuwen Liu, Jiang Bian, Zhe He

**Author notes:** Corresponding author: Zhe He.

## Abstract

Acute myocardial infarction poses significant health risks and financial burden on healthcare and families. Prediction of mortality risk among AMI patients using rich electronic health record (EHR) data can potentially save lives and healthcare costs. Nevertheless, EHR-based prediction models usually use a missing data imputation method without considering its impact on the performance and interpretability of the model, hampering its real-world applicability in the healthcare setting. This study examines the impact of different methods for imputing missing values in EHR data on both the performance and the interpretations of predictive models. Our results showed that a small standard deviation in root mean squared error across different runs of an imputation method does not necessarily imply a small standard deviation in the prediction models’ performance and interpretation. We also showed that the level of missingness and the imputation method used can have a significant impact on the interpretation of the models.

## Introduction

Cardiovascular diseases (CVDs) remain the leading cause of death worldwide ^1^ and account for 1 in 3 deaths of adults in the United States every year.^2^ CVDs cause a heavy toll on the health and economy all over the world.^3^ Among various CVDs, acute myocardial infarction (AMI) is the most severe form of coronary artery disease and a fatal CVD responsible for the death of millions of people annually around the world.^4^ Thus, prediction of mortality risk among AMI patients is important for early interventions or advising preventive strategies to high risk patients, which will save lives and costs.

The wide adoption of electronic health records (EHR) systems in the United States is the result of a series of government initiatives^5^ and led to a large amount of clinical data accumulated in digital forms.^6^ EHR data is a rich source of patient information for predictive analysis in healthcare.^7^ Predictive analysis in healthcare and clinical decision-support is not a new topic.^8^ Nevertheless, in recent years, there is an increasing demand of using routinely collected real-world data (RWD) such as EHRs, administrative claims, and billing data to generate real-world evidence (RWE) that informs regulatory decisions and clinical care.^9^ On the other hand, the emergence and efficient implementation^10^ of state-of-the-art machine learning,^11^ especially deep learning methods,^12^ as well as increasingly powerful computing infrastructure make predictive analysis using EHR data more possible than ever.

Nevertheless, using EHR data for predictive analysis with machine learning and deep learning methods is still challenging. One major issue is the quality of EHR data due to incompleteness.^13^ The existence of missing values in EHR data is multi-fold, including human errors such as the lack of collection (e.g., the medical expert did not perform an evaluation) and the lack of documentation (e.g., the medical expert did not document an evaluation result).^14^ Thus, a significant body of literature has attempted to approach the missing value issue by imputation, rather than eliminating records with missing data entirely (i.e., as it reduces the sample size).^15^ Mean or median value imputation is a common approach for imputing missing values in EHR data mainly due to its ease of implementation.^16^ Researchers have also proposed multiple imputation by chained equations (MICE)^17^ or its variations to deal with missing values in EHR data.^14^ There are also machine-learning-based imputation methods such as MissForest^18^ and K-nearest neighbors (KNN)-based imputation.^19^ A few recent studies have also used deep learning methods such as generative adversarial networks (GANs)^20^ and autoencoders for missing value imputation in EHR data.^19^

Depending on the reason for a value to be missing in a dataset, there are three main categories of missingness mechanism, including missing completely at random (MCAR), missing at random (MAR), and not missing at random (NMAR).^21^ Characterizing the missingness mechanism in EHR data can be an indicator of choosing an appropriate imputation method. This impact has been explored previously^19^ and is not within the scope of this study.

On the other hand, predictive analysis using complex machine learning methods (e.g., deep learning), which yield superior prediction accuracy, usually result in black-box models that are not interpretable by the end-users. Despite their promising performance, using such complex predictive models in healthcare and clinical decision-making process is quite challenging. Medical professionals need to understand the rationale behind the predictive models’ predictions.^22^ Thus, they prefer less complex models such as logistic regression for clinical decision-making.^12^ Researchers in the field have taken different approaches to address the interpretability of machine learning models, for instance, feature interaction and importance, attention mechanism, data dimensionality reduction, knowledge distillation and rule extraction.^23^ Nevertheless, there are still some fundamental issues that need to be addressed such as fidelity of the post-hoc interpretation methods to the reference model, evaluation of the interpretation methods, and design biases due to focusing on the intuition of researchers rather than real end-users’ (medical professionals in this context) needs. For more reading on this we refer the interested audience to a recent systematic review on the explainable AI models using EHR data.^23^

For example, in a logistic regression model for binary outcome, the coefficients of the features (predictors) can be readily transformed into odds ratios and can be easily understood as feature importance. Nevertheless, as these coefficients are estimated from the input data, when the data points were replaced with different imputation techniques, the extent to which missing data points are extrapolated has certain impact on the interpretation of these coefficients. Further, in real-world applications of EHR and in the absence of a complete dataset (knowing the ground truth), it would not be straightforward to choose the best imputation method.^19^ Although missingness has been recognized as a major data quality issue of EHR,^24^ the impact of imputation methods on the interpretations of machine learning models has not been well explored and warrants more investigation. Thus, in this study, we examine (1) the performance of imputation methods on missing values in EHR data, (2) the impact of different imputation methods on the performance, and (3) the interpretations of predictive models, using all-cause mortality among AMI patients as a case study.

## Methods

Figure 1 illustrates the overall workflow of this study. First, we identified patients with AMI from the Medical Information Mart for Intensive Care (MIMIC-III) dataset and created a complete dataset without missing values as the baseline. We then introduced different levels of missingness (i.e., from 10% to 50%) through simulations. Then, we applied different statistical and machine learning-based imputation methods including mean (mode for two categorical variables), MICE, MissForest, and a KNN-based method, as well as Generative Adversarial Imputation Networks (GAIN)^20^ - a novel imputation method based on neural networks. Then, we compared these imputation methods’ performance in terms of root mean square error (RMSE), which measures the difference between the imputed values (in the datasets with missing value that were imputed) and the actual values (in the complete data set).^30^ Further, we built shallow machine learning models that are intrinsically interpretable and preferred by medical experts, such as logistic regression, linear support vector machine (SVM), and decision tree. We compared the performance of the models that were based on the imputed datasets in terms of area under the receiver operator characteristic curve (AUROC) against the performance of the reference model (i.e., the model based on the complete data set). For simplicity, we refer to the former as the “imputed-data models” and the latter as the “reference model” throughout the paper. Finally, we compared the feature importance derived from the imputed-data models against the feature importance derived from the corresponding reference model using Pearson correlation analysis. When the underlying data changes (i.e., from complete dataset to one with imputed values), the resulting model characteristics might change as well as the produced feature importance. Since we observed variance in the performance of the shallow models, we employed the DeepConsensus^25^ algorithm to investigate the impact of consensus mechanism among deep models on reducing performance variance. We elaborate more on this in the “Predictive Modeling” subsection.

**Figure 1.**
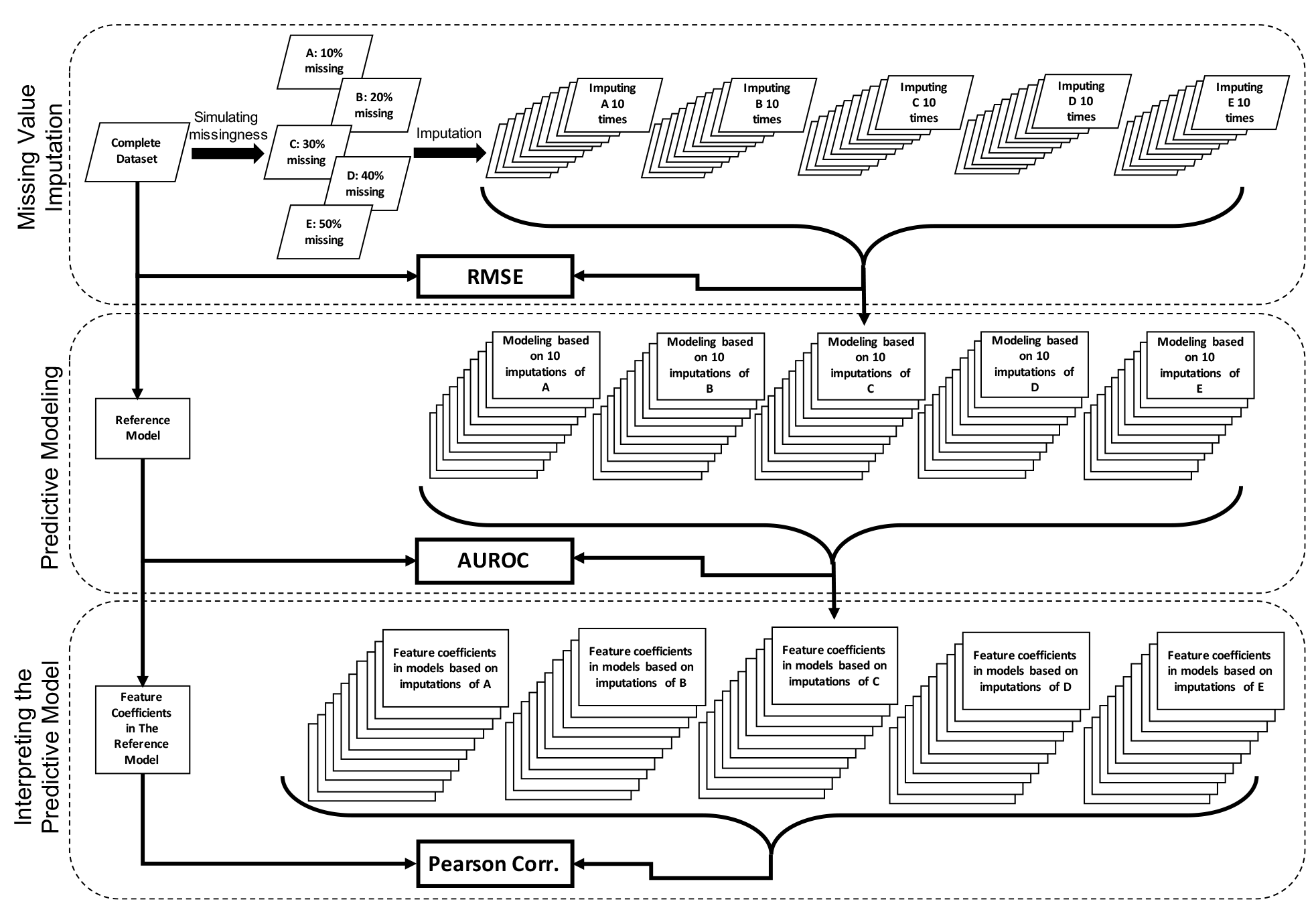
The workflow of this study. For simplicity, this figure only illustrates the workflow for one imputation method and one prediction model.

### Data Source

The Medical Information Mart for Intensive Care (MIMIC-III) database is an integration of de-identified and comprehensive EHR data of patients admitted to the Beth Israel Deaconess Medical Center in Boston, Massachusetts.^26^ This dataset is freely available and contains patient information spanning over more than a decade. Considering the importance of studying the all-cause mortality risk among patients with cardiovascular diseases, especially AMI, in this study, we focused on patients with AMI and post myocardial syndrome (PMS). The International Classification of Diseases, 9^th^ revision, Clinical Modification (ICD-9-CM) codes considered for this study are 410.0 to 411.0. For each admission, we aggregated the laboratory and chart values and considered the average value of each of the 19 numerical features. We also considered two categorical features with few missing values including gender and initial emergency room diagnosis of AMI. Since our purpose for this study was to examine how the ranking of feature importance would change under different imputation methods and level of missingness, we did not consider the longitudinal dimension of features. In order to build a reference dataset with no missing values, we excluded features with more than 50% missing values and applied listwise deletion to the rest of the original dataset. The resulting complete dataset has 3054 observations and 21 features. The binary outcome was all-cause mortality within one year of admission. Features description and summary statistics are reported in Table 1.

**Table 1.** Summary statistics of the variables in the reference complete dataset with 3054 instances.

| Variable Name | Description | Variable Type | Min | Max | Median | Mean | Standard Deviation |
| --- | --- | --- | --- | --- | --- | --- | --- |
| Diastolic BP | Diastolic blood pressure | Numerical | 18.31 | 134.69 | 51.2 | 52.2 | 11.28 |
| Systolic BP | Systolic blood pressure | Numerical | 37.97 | 484.12 | 104.96 | 105.44 | 22.74 |
| Heart Rate | Heart rate | Numerical | 42 | 139.48 | 83.81 | 84.13 | 11.97 |
| Resp Rate | Respiratory rate | Numerical | 9 | 42.7 | 19.33 | 19.62 | 3.28 |
| Bicarbonate | Bicarbonate | Numerical | 8 | 41.88 | 24.92 | 24.61 | 3.65 |
| Calcium | Calcium | Numerical | 5.6 | 13.95 | 8.43 | 8.44 | 0.59 |
| Chloride | Chloride | Numerical | 80.42 | 125.61 | 104 | 104.01 | 4.55 |
| Potassium | Potassium | Numerical | 1.87 | 6.9 | 4.14 | 4.18 | 0.36 |
| Sodium | Sodium | Numerical | 118.18 | 158.5 | 138.72 | 138.67 | 3.47 |
| Glucose | Glucose | Numerical | 65.67 | 543 | 131.76 | 141.25 | 39.93 |
| Hematocrit | Hematocrit | Numerical | 21.11 | 50.61 | 30.97 | 31.53 | 3.6 |
| Hemoglobin | Hemoglobin | Numerical | 6.4 | 16.27 | 10.43 | 10.63 | 1.34 |
| WBC | White blood cell count | Numerical | 0.45 | 107.68 | 10.93 | 11.75 | 5.19 |
| ALT | Alanine aminotransferase | Numerical | 2 | 5509 | 30.33 | 89.71 | 270.59 |
| AST | Aspartate aminotransferase | Numerical | 2 | 13511.7 | 46 | 142.36 | 486.22 |
| ALP | Alkaline phosphatase | Numerical | 19 | 1147.92 | 80 | 102.61 | 83.6 |
| Albumin | Albumin | Numerical | 1.2 | 5 | 3.2 | 3.2 | 0.6 |
| Bilirubin | Bilirubin | Numerical | 0.1 | 31.14 | 0.6 | 0.97 | 1.77 |
| Admit Age | Age at admission | Numerical | 21.22 | 97.52 | 72.55 | 70.89 | 12.96 |
| InitialERDiagnosisMI (0 = No, 1 = Yes) | Initial emergency room diagnosis was AMI or rule out AMI (#0s = 2050, #1s = 1004) | Categorical | Not applicable |  |  |  |  |
| Gender (0 = Female, 1 = Male) | Gender (#0s = 1202, #1s = 1852) | Categorical |  |  |  |  |  |

### Missingness Mechanisms

In this study, our focus is not on studying missingness mechanisms, rather it is on exploring the impact of imputation methods on models’ performance, and more importantly, the derived interpretations. We acknowledge that MCAR is not the only possible missingness mechanism in RWD such as EHRs. To give an example, medical professionals might less likely order fasting glucose tests for healthier patients in comparison to those with risk factors of diabetes. Thus, for healthier patients, in this case, there might be more missing glucose values. However, covering MAR and NMAR missingness mechanisms is out of the scope of this paper. Considering all possible combinations of missingness mechanisms with other criteria experimented in this paper would exponentially expand the scope of the paper and increase the complexity of reporting. We simulated random missingness on the complete dataset by removing random 10%, 20%, 30%, 40%, and 50% of values. Any value in the data was as likely to be missing as any other value. Thus, the missingness mechanism in this study was MCAR.

### Imputation Methods

In this study, we evaluated five different imputation methods, including (1) mean value imputation, (2) MICE, (3) K-nearest neighbors (KNN)-based, (4) MissForest, and (5) GAIN. For implementation purposes, we used available packages in R to implement methods (1) to (4). The implementation code of GAIN in Python is made available by its authors on GitHub (https://github.com/jsyoon0823/GAIN). The performance of different imputation methods was compared using RMSE. The details of these methods are described as follows.

**Mean value imputation:** The easiest to implement and most conventional approach to impute missing values in EHR data is mean value imputation. However, this simplicity might result in ignoring the underlying statistical information in data and introduce unintentional biases in the subsequent analyses.^14^

**MICE:** MICE is one of the most popular methods for imputing missing values in EHR data. The main reason resides in its ability to impute different types of variables that might be present in the EHR data. Using MICE, each variable with missing observations is regressed on all the remaining variables in the dataset. The missing values are replaced with the predicted value, and this imputation process is repeated sequentially until all missing values are imputed.

**KNN-based:** KNN is a machine learning method that can be used for imputing missing values in EHR data.^19^ In this approach, missing values are replaced with the mean value of *k* most similar complete observations. A distance function (e.g., Euclidean) is used to measure this similarity.

**MissForest:** MissForest is a promising imputation method for missing values in EHR data.^18^ In this method, first, mean imputation (or any other imputation method) is performed as an initial guess for the missing values. Then, variables in the dataset are sorted based on the amount of missing values they have with the one with fewest missing values ordered first. Further, for each variable x, a random forest ^27^ model is fitted on all other variables’ observed values and the outcome variable being the observed values of variable *x*. Then, the trained model is used to predict the missing values of x. This process is repeated until a stopping criterion is met.

**GAIN:** Recently, GAIN, a neural network-based imputation method was introduced for missing value imputation. This imputation method is based on the generative adversarial networks (GAN)^20^ framework. In the framework, corresponding to a minimax two-player game, two models are trained simultaneously, a generative model and a discriminative model. The generative model captures the data distribution while the discriminative model estimates the probability of a sample being from the training data or from the generative model. The objective of the generative model is to make the discriminative model make more mistakes. In GAN, the generative and discriminative models are defined based on multilayer perceptron (feedforward neural networks). GAIN is an imputing GAN framework in which the goal of the generative model is to accurately impute the missing values in data, while the goal of the discriminative model is to predict the probability of a value being from the original dataset or from the generative model (observed or imputed component). The objective of the discriminative model in GAIN is to minimize the error loss (on guessing if the elements in the generative model’s output are produced by the generative model or from the original data) while the generative model’s goal is to maximize discriminative model’s mistakes. The authors of GAIN have reported superior imputation performance of GAIN in comparison to autoencoders and other statistical and conventional machine learning-based imputation methods. For more information on GAIN, we refer the interested audience to the original paper.^20^

### Predictive Modeling

To compare the prediction performance and feature importance ranking of different prediction models with different level of missingness, we performed predictive analysis using three popular shallow machine learning methods in predictive modeling with EHR data,^28^ namely logistic regression, SVM, and decision tree. To keep consistency, the same configurations (default) were used across all models based on the imputed datasets.^29^ Hypothetically, an effective imputation method should approximate values that are very close to the original values of the missing data. Thus, to understand how the performance and interpretations change under different imputation methods, we kept the configurations consistent from the reference model to the imputed-data models with gradually increasing amount of missing values. Further, we captured feature coefficients (importance) in each imputed-data model to compare to the same in the reference model of its own kind. For comparison, we used Pearson correlation coefficients. A higher correlation means closer results from the imputed-data model (in terms of feature importance) to the reference model based on the complete dataset. Also, we built a deep learning model (i.e., DeepConsensus) to investigate if it can reduce the variance of the models’ classification performance. The binary prediction task was patient all-cause mortality within one year after admission. The dataset was divided to separate training and testing sets at the ratio of 0.9 to 0.1 respectively. The dataset is imbalanced with 65 (negative class) to 35 (positive class) ratio. For implementation purposes we used Python programming language with Tensorflow, NumPy, Pandas, and Sklearn packages. We give a brief description of DeepConsensus in the following.

**DeepConsensus:** The main idea behind DeepConsensus is that since different deep neural networks tend to classify training samples accurately, they generate similar linear regions. Thus, these models should behave similarly on classifying training samples. Such behavior enables multiple models to agree with each other on classifying valid inputs and filtering out adversarial examples, while individual models are sensitive to those examples. Using consensus among different models helps to capture the underlying structure of data. It is shown that consensus helps to differentiate extrinsically classified samples (i.e., classified under extrinsic factors such as randomness of weight initialization) from consistently classified samples (i.e., samples that are classified in the same class with high probability by multiple models). Thus, such consensus mechanism among multiple models can reduce the variance caused by extrinsic factors. The effectiveness of this method is demonstrated in the reference paper.^25^ Note that in this study, DeepConsensus was only employed to investigate the impact of consensus mechanism on the variance of the classification performance. In terms of model interpretations, to keep consistency across all experiments, we only focused on less complex machine learning models that are intrinsically interpretable (DeepConsensus utilizes a posthoc interpretation approach).^25^ Five individual deep models were trained using the same 90% of the records and evaluated on the remining 10%. All models consisted of 4 hidden dense layers. The implementation details of these models are provided in Table 2.

**Table 2.** Implementation detail of five individual models in DeepConsensus.

| **Specification** | **Model 1** | **Model 2** | **Model 3** | **Model 4** | **Model 5** |
| --- | --- | --- | --- | --- | --- |
| **#Neurons in each layer** | 105 | 130 | 130 | 140 | 105 |
| **Activation function** | ReLU | Tanh | ReLU | SeLU | ReLU |
| **Optimization** | Adagrad | Adamax | RMSprop | Adam | Adagrad |
| **Bias** | Random Uniform | Zeros | Constant | Zeros | Random Uniform |
| **Weights** | Random Normal | Glorot Uniform | Random Normal | Random Normal | Random Normal |

## Results

### Imputation Performance

We compared five different imputation methods including MICE, MissForest, KNN-based, Mean (mode for categorical variables), and GAIN, on 10% to 50% missing datasets. We ran each experiment 10 times and computed the average RMSE along with its standard deviation. The results are reported in Table 3.

**Table 3.** Average RMSE from different imputation methods on 10% to 50% missing data (10 runs).

| Missingness level<br>Imputation Method | 10% | 20% | 30% | 40% | 50% |
| --- | --- | --- | --- | --- | --- |
| MICE | 0.2254±0.0025 | 0.2249±0.0020 | 0.2292±0.0013 | 0.2343±0.0014 | 0.2325±0.0014 |
| MissForest | 0.1935±0.0017 | 0.1950±0.0013 | 0.1997±0.0018 | 0.2051±0.0018 | 0.2062±0.0011 |
| KNN-based | 0.21447 | 0.2219 | 0.2241 | 0.2267 | 0.2228 |
| Mean/Mode | 0.2038 | 0.2046 | 0.2052 | 0.2066 | **0.2059** |
| GAIN | **0.1757±0.0049** | **0.1763±0.0064** | **0.1838±0.0039** | **0.1963±0.0114** | 0.2088±0.0100 |

Averaging the RMSE of different imputation methods across all different levels of missing values, GAIN showed the best performance. MissForest was the second-best performing imputation method following GAIN showing only 0.012 difference in RMSE on average. Mean came next, showing quite monotonic behavior across all datasets with different amount of missing values. KNN-based and MICE showed similar performance reporting the highest RSME (worst performance).

### Prediction Performance

Next, to narrow down the required experiments, we focused on the datasets imputed with the best performing imputation methods in terms of RMSE: GAIN and MissForest. First, we built the reference models that served as the benchmark for our comparisons. Then, we built models based on datasets with varying percentage of missingness that were imputed using GAIN and MissForest (through 300 experiments = 5 levels of missingness * 2 imputation methods* 3 ML methods * 10 runs each). The performance of these models based on GAIN imputed datasets and MissForest imputed datasets in terms of AUROC is depicted in Figure 2. AUROC is a classification performance measure that illustrates how models perform in terms of discriminating between the classes. The performance of SVM and logistic regression models based on MissForest imputed datasets (Figure 2) showed lower variance in comparison to the same models based on GAIN imputed datasets. The variance in decision tree models based on the datasets imputed by both GAIN and MissForest is relatively high, making those models’ performance unstable even under low levels of missingness.

**Figure 2.**
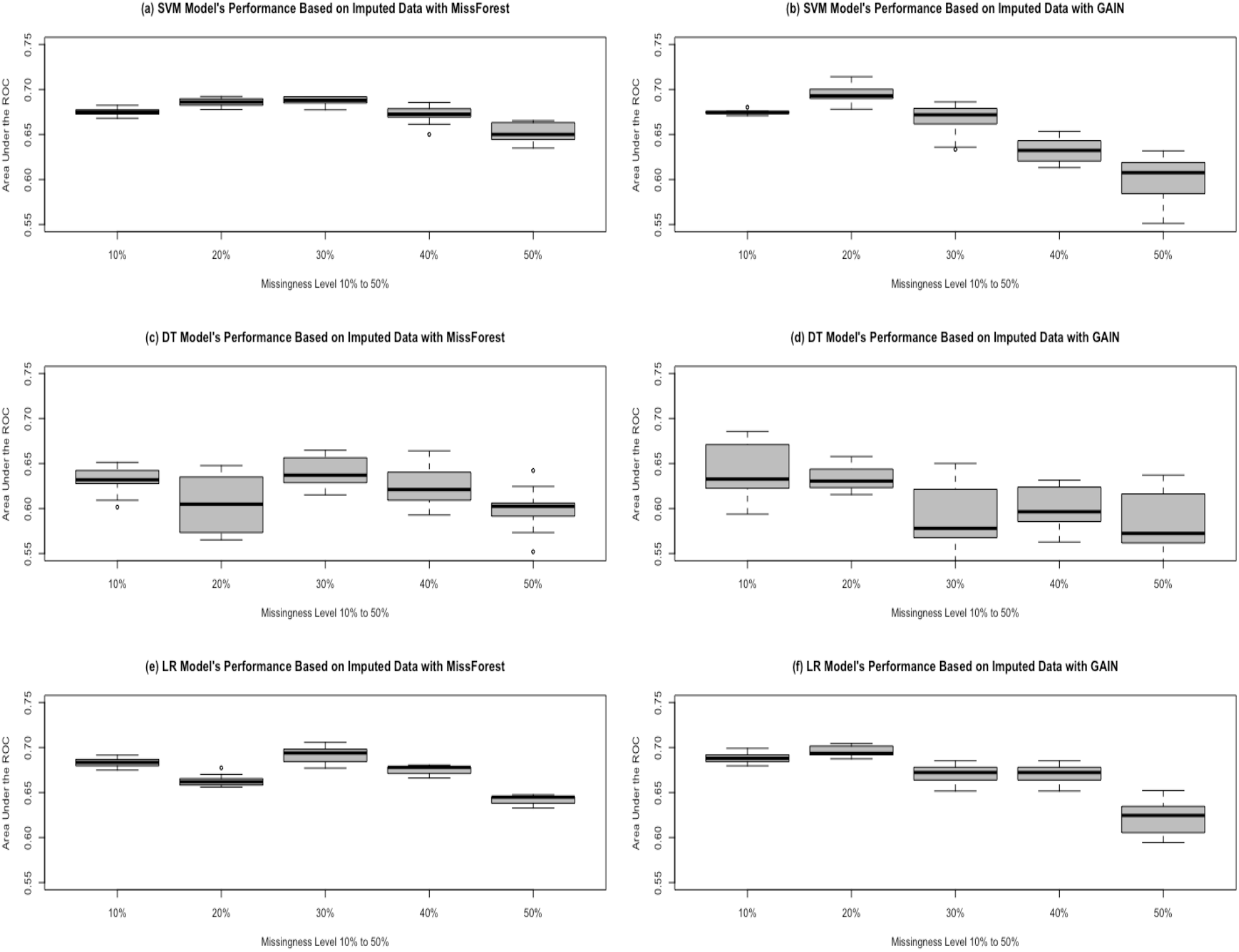
Comparing the performance of models that are built using (a) support vector machine (SVM) based on the imputed datasets with MissForest, (b) SVM based on the imputed datasets with GAIN, (c) decision tree (DT) based on the imputed datasets with MissForest, (d) DT based on the imputed datasets with GAIN, (e) logistic regression (LR) based on the imputed datasets with MissForest, (f) LR based on the imputed datasets with GAIN, under gradually increasing missingness level. The performance is reported on area under the receiver operating characteristic curve (AUROC).

By increasing the number of missing values in the datasets from 10% to 50%, models based on the datasets imputed by MissForest showed a more stable behavior in comparison to GAIN across all models. A closer look at SVM performance (based on AUROC) from 10% to 50% missing values imputed by MissForest showed an increase of standard deviation from 0.003 to 0.010 with an average of 0.006. This measure was 0.002 to 0.025 for GAIN with an average of 0.014. Also, a big jump in standard deviation was not observed in case of MissForest until 40% of missingness. This jump occurred at 20% of missingness in case of GAIN. The standard deviation of AUROC of multiple runs for logistic regression models based on GAIN showed an increasing trend from 0.005 to 0.019 with the average of 0.010. However, in case of MissForest, the trend is increasing from 10% to 30% and then decreasing from 30% to 50% with an average of 0.006. We hypothesized that the variance among models on the same missing level of missing data that is imputed with the same method 10 times can be reduced by using the consensus mechanism among multiple deep models. Applying DeepConsensus on the complete dataset (baseline) showed a significant increase of performance as expected: from 0.721 AUROC in logistic regression and 0.713 in SVM to 0.809 in DeepConsensus. Further, we narrowed down our experiments to 10 datasets that were the result of imputing 10% missingness (lowest missing rate) 10 times with MissForest (imputation method with less variance on machine learning models). The average AUROC across 10 experiments showed an increase of performance to 0.790 (from 0.675 in SVM and 0.683 in logistic regression). However, the variance in performance still persists at 0.0344.

### Feature Importance Ranking Comparison

The Pearson correlation of feature importance resulted from each of the imputed-data models with that of the corresponding reference models is reported in Table 4. The goal of this analysis was to investigate how the reported feature importance would change under different levels of missingness and under different imputation techniques. In other words, how similar the feature importance in each imputed-data model is to the corresponding reference model. These results showed a generally lower performance for decision tree models across all datasets. Focusing on SVM and logistic regression with higher performance, averaging coefficients as a result of 10 runs for each level of missingness showed a correlation coefficient of more than 0.99 with statistically significant results on the imputed datasets of 10% missing value with GAIN and MissForest. However, this trend on models based on an increasing level of missingness on average showed a decreasing correlation with the reference model across all machine learning methods and imputation methods. A further principal component analysis (PCA) on the complete dataset showed that a linear model based on 30% missing values could capture between 90.130% (14 features) and 92.824% (15 features) of the statistical information in this dataset.

**Table 4.** Pearson correlation coefficients and p-values of feature importance comparison between the imputed-data models and the reference models.

| Machine Learning method | Imputation method | Missingness % | Pearson correlation coefficient | p-value | Imputation method | Missingness % | Pearson correlation coefficient | p-value |
| --- | --- | --- | --- | --- | --- | --- | --- | --- |
| Decision Tree | GAIN | 10% | 0.966 | 1.24E-12 | MissForest | 10% | 0.944 | 1.23E-10 |
|  |  | 20% | 0.959 | 7.09E-12 |  | 20% | 0.914 | 6.56E-09 |
|  |  | 30% | 0.917 | 4.84E-09 |  | 30% | 0.906 | 1.49E-08 |
|  |  | 40% | 0.842 | 1.64E-06 |  | 40% | 0.874 | 2.24E-07 |
|  |  | 50% | 0.903 | 2.05E-08 |  | 50% | 0.770 | 4.34E-05 |
| SVM |  | 10% | 0.992 | 8.57E-19 |  | 10% | 0.994 | 7.71E-20 |
|  |  | 20% | 0.986 | 1.80E-16 |  | 20% | 0.983 | 1.12E-15 |
|  |  | 30% | 0.949 | 5.54E-11 |  | 30% | 0.985 | 5.31E-16 |
|  |  | 40% | 0.834 | 2.51E-06 |  | 40% | 0.975 | 5.09E-14 |
|  |  | 50% | 0.793 | 1.73E-05 |  | 50% | 0.911 | 8.83E-09 |
| Logistic Regression |  | 10% | 0.995 | 6.80E-21 |  | 10% | 0.996 | 2.56E-22 |
|  |  | 20% | 0.988 | 5.14E-17 |  | 20% | 0.987 | 1.28E-16 |
|  |  | 30% | 0.962 | 3.62E-12 |  | 30% | 0.985 | 4.24E-16 |
|  |  | 40% | 0.895 | 4.00E-08 |  | 40% | 0.978 | 1.53E-14 |
|  |  | 50% | 0.854 | 8.39E-07 |  | 50% | 0.958 | 7.44E-12 |

## Discussion

Comparing the imputation methods’ RMSE reported in Table 3 implies that (1) GAIN performs better than MissForest, and (2) the standard deviation between different runs of the same method on the same dataset with missing values is small. However, our experiments confirmed that choosing the best imputation method may not always be a straightforward process. Although GAIN surpassed all imputation methods in terms of RMSE on all datasets, MissForest imputation yielded more stable results (smaller standard deviation on average) at the presence of gradually increasing number of missing values. Also, comparing the performance of models based on datasets with different percentage of missingness reveals the fact that higher performance does not necessarily indicate more similar interpretations to the reference model. We observed that on average a relatively small standard deviation of RMSE across all levels of missingness yielded a bigger standard deviation in models’ performance and a lower correlation of feature importance between the reference models and the imputed-data models. Also, the dilemma of bias/variance is well understood regarding neural networks which are hyperparameterized.^31^ Training neural networks requires a larger number of training samples to achieve acceptable performance and less variance. Thus, although using consensus of deep models did not resolve the issue of variance in this study, we hypothesize that using a bigger dataset (with more samples and more features) could potentially yield a more stable consensus of deep learning models and result in less variance.

These observations might not be generalizable to other datasets or imputation methods. There is no universally optimal approach for missing data imputation or predictive modeling using EHR data. However, these experiments showed that the way we approach missing values in EHR data impacts not only the model performance but also the interpretations of the models’ predictions. In real-world predictive analysis of EHR data, it is usually not possible to obtain a dataset with no missing values. However, in cases where the interpretations of predictive models matter, in order to choose the best imputation method, just relying on RMSE or model performance measures may not be sufficient. In these cases, we suggest running extensive experiments on a smaller complete-case version of the dataset first, evaluate the impact of different imputation methods on the interpretations in comparison to the complete-case, and then apply the best performing method on the original dataset with missing values. In cases where it is not possible to have the complete-case dataset, researchers should be aware of this potential impact, use different imputation methods for predictive modeling, and discuss the resulting interpretations with medical experts or compare to the medical knowledge when choosing the imputation method that yields the most reasonable interpretations. Also, more in-depth analyses of data with methods such as PCA can be used to investigate the redundancy in datasets and determine the maximal allowed missing value rate.

### Limitations and Future Opportunities

A potential limitation of this study was the relatively small and imbalanced dataset (65:35). Although the findings in this study are robust, future studies could be done on datasets with more balance and more samples to investigate how these results would change. We acknowledge that there are different missingness mechanisms, missing value imputation methods, predictive modeling approaches, and interpretability enhancement techniques that are not covered in this study. However, implementing all the possible combinations of these criteria is expensive. The main intention behind this case study was to demonstrate the potential impact of missing value imputation on derived interpretations of predictive models. Thus, we encourage future in-depth theoretical and applied studies on this topic. We believe the interpretability issue in predictive modeling is of same importance, if not more, as performance when it comes to medical applications. Also, the interpretability enhancement of predictive models based on longitudinal EHR data is inherently challenging and has not been extensively explored. However, we believe investigating the potential impact of missing value imputation in longitudinal EHR data and its interpretations is an important future direction and deserves more investigation.

## Conclusions

In this study, we simulated 5 levels of missingness (10% to 50%) on a complete EHR dataset of 21 features for 3054 patients with AMI from MIMIC-III database. We examined different statistical and machine learning-based imputation methods such as mean, MICE, MissForest, and KNN-based, as well as GAIN-a novel imputation method based on GAN. Our experiments showed that GAIN and MissForest yielded best performance in terms of RMSE and small standard deviations across all levels of missingness. However, further predictive modeling based on each of these datasets revealed the fact that the variance in their performance (in terms of AUROC) gradually grows with more missingness. Also, Pearson correlation analysis showed that the similarity of feature importance of models based on the imputed datasets to the feature importance of baseline models gradually decreases, a trend that could not initially be inferred by just looking at the performance of imputation and predictive modeling in terms of RMSE and AUROC respectively.

## Data Availability

The data are available upon request.

## Acknowledgments

This study was supported in part by the National Institute on Aging (NIA) of the National Institutes of Health (NIH) under Award Number R21AG061431; and the University of Florida Clinical and Translational Science Institute, which is supported in part by the NIH National Center for Advancing Translational Sciences under award number UL1TR001427. The content is solely the responsibility of the authors and does not necessarily represent the official views of the NIH.

## Notes

### Competing Interest Statement

The authors have declared no competing interest.

### Author Declarations

IRB is not applicable due to the use of de-identified public dataset.

## References

1. al-Aiad A, Duwairi R, Fraihat M. Survey: Deep Learning Concepts and Techniques for Electronic Health Record. In: 2018 IEEE/ACS 15th International Conference on Computer Systems and Applications (AICCSA). 2018. p. 1-5.

2. Curry SJ, Krist AH, Owens DK, Barry MJ, Caughey AB, Davidson KW, et al. Risk Assessment for Cardiovascular Disease With Nontraditional Risk Factors: US Preventive Services Task Force Recommendation Statement. JAMA. 2018 Jul 17;320(3):272–80.

3. Ford ES, Capewell S. Coronary Heart Disease Mortality Among Young Adults in the U.S. From 1980 Through 2002: Concealed Leveling of Mortality Rates. Journal of the American College of Cardiology. 2007 Nov 27;50(22):2128–32.

4. Reed GW, Rossi JE, Cannon CP. Acute myocardial infarction. The Lancet. 2017 Jan 14;389(10065):197–210.

5. Blumenthal D. Implementation of the Federal Health Information Technology Initiative. New England Journal of Medicine. 2011 Dec 22;365(25):2426–31.

6. ANSI I. ISO/DTR 20514: Health informatics-electronic health record—definition, scope and context. ISO; 2005.

7. Milenkovic MJ, Vukmirovic A, Milenkovic D. Big data analytics in the health sector: challenges and potentials. Management: Journal of Sustainable Business and Management Solutions in Emerging Economies. 2019;24(1):23–33.

8. Parikh RB, Kakad M, Bates DW. Integrating Predictive Analytics Into High-Value Care: The Dawn of Precision Delivery. JAMA. 2016 Feb 16;315(7):651–2.

9. Sherman RE, Anderson SA, Dal Pan GJ, Gray GW, Gross T, Hunter NL, et al. Real-World Evidence — What Is It and What Can It Tell Us? New England Journal of Medicine. 2016 Dec 8;375(23):2293–7.

10. Erickson BJ, Korfiatis P, Akkus Z, Kline T, Philbrick K. Toolkits and Libraries for Deep Learning. J Digit Imaging. 2017 Aug 1;30(4):400–5.

11. Steele AJ, Denaxas SC, Shah AD, Hemingway H, Luscombe NM. Machine learning models in electronic health records can outperform conventional survival models for predicting patient mortality in coronary artery disease. PLoS One [Internet]. 2018 Aug 31 [cited 2020 Feb 20];13(8). Available from: https://www.ncbi.nlm.nih.gov/pmc/articles/PMC6118376/

12. Shickel B, Tighe PJ, Bihorac A, Rashidi P. Deep EHR: A Survey of Recent Advances in Deep Learning Techniques for Electronic Health Record (EHR) Analysis. IEEE Journal of Biomedical and Health Informatics. 2018 Sep;22(5):1589–604.

13. Botsis T, Hartvigsen G, Chen F, Weng C. Secondary Use of EHR: Data Quality Issues and Informatics Opportunities. Summit on Translat Bioinforma. 2010 Mar 1;2010:1-5.

14. Wells BJ, Chagin KM, Nowacki AS, Kattan MW. Strategies for handling missing data in electronic health record derived data. EGEMS (Wash DC). 2013;1(3):1035.

15. Little RJ, Rubin DB. Statistical analysis with missing data. Vol. 793. John Wiley & Sons; 2019.

16. Salgado CM, Azevedo C, Proença H, Vieira SM. Missing Data. In: MIT Critical Data, editor. Secondary Analysis of Electronic Health Records [Internet]. Cham: Springer International Publishing; 2016 [cited 2020 Feb 20]. p. 143-62. Available from: https://doi.org/10.1007/978-3-319-43742-2_13

17. Buuren S van, Groothuis-Oudshoorn K. mice: Multivariate imputation by chained equations in R. Journal of statistical software. 2010;1-68.

18. Stekhoven DJ, Bühlmann P. MissForest—non-parametric missing value imputation for mixed-type data. Bioinformatics. 2012 Jan 1;28(1):112–8.

19. Beaulieu-Jones BK, Lavage DR, Snyder JW, Moore JH, Pendergrass SA, Bauer CR. Characterizing and managing missing structured data in electronic health records: data analysis. JMIR medical informatics. 2018;6(1):e11.

20. Yoon J, Jordon J, Schaar M. GAIN: Missing Data Imputation using Generative Adversarial Nets. In: International Conference on Machine Learning [Internet]. 2018 [cited 2020 Feb 27]. p. 5689-98. Available from: http://proceedings.mlr.press/v80/yoon18a.html

21. Scheffer J. Dealing with missing data. 2002 [cited 2020 Feb 28]; Available from: https://mro.massey.ac.nz/handle/10179/4355

22. Ahmad MA, Eckert C, Teredesai A. Interpretable Machine Learning in Healthcare. In: Proceedings of the 2018 ACM International Conference on Bioinformatics, Computational Biology, and Health Informatics [Internet]. Washington, DC, USA: Association for Computing Machinery; 2018 [cited 2020 Feb 21]. p. 559-560. (BCB ’18). Available from: https://doi.org/10.1145/3233547.3233667

23. Payrovnaziri SN, Chen Z, Rengifo-Moreno P, Miller T, Bian J, Chen JH, et al. Explainable artificial intelligence models using real-world electronic health record data: a systematic scoping review. J Am Med Inform Assoc. 2020 Jul 1;27(7):1173–85.

24. Weiskopf NG, Weng C. Methods and dimensions of electronic health record data quality assessment: enabling reuse for clinical research. J Am Med Inform Assoc. 2013;20(1):144–51.

25. Salman S, Payrovnaziri SN, Liu X, Rengifo-Moreno P, He Z. Consensus-based Interpretable Deep Neural Networks with Application to Mortality Prediction. arXiv:190505849 [cs, stat] [Internet]. 2019 Sep 11 [cited 2020 Mar 11]; Available from: http://arxiv.org/abs/1905.05849

26. Johnson AEW, Pollard TJ, Shen L, Lehman LH, Feng M, Ghassemi M, et al. MIMIC-III, a freely accessible critical care database. Scientific Data. 2016 May 24;3(1):1–9.

27. Breiman L. Random Forests. Machine Learning. 2001 Oct 1;45(1):5–32.

28. Taslimitehrani V, Dong G, Pereira NL, Panahiazar M, Pathak J. Developing EHR-driven heart failure risk prediction models using CPXR(Log) with the probabilistic loss function. Journal of Biomedical Informatics. 2016 Apr 1;60:260-9.

29. Farhangfar A, Kurgan L, Dy J. Impact of imputation of missing values on classification error for discrete data. Pattern Recogn. 2008 Dec 1;41(12):3692–3705.

30. Duy Le T, Beuran R, Tan Y. Comparison of the Most Influential Missing Data Imputation Algorithms for Healthcare. In: 2018 10th International Conference on Knowledge and Systems Engineering (KSE). 2018. p. 247-51.

31. Geman S, Bienenstock E, Doursat R. Neural networks and the bias/variance dilemma. Neural computation. 1992;4(1):1–58.

